# Physical activity and posture profile of a South African cohort of middle-aged men and women as determined by integrated hip and thigh accelerometry

**DOI:** 10.1101/2021.10.22.21265362

**Authors:** Lisa K. Micklesfield, Kate Westgate, Antonia Smith, Clement Kufe, Amy E. Mendham, Tim Lindsay, Katrien Wijndaele, Julia H. Goedecke, Soren Brage

## Abstract

**Background:** Physical activity and sedentary behaviour are central to public health recommendations and highlight the need for precise measurement. Descriptive studies of objectively measured physical activity behaviours in African populations are rare. We aimed to develop a method of combining the signals from hip and thigh accelerometers to quantify physical behaviours and describe these by socio-demographic factors in a population of middle-aged men and women from urban South Africa.

**Methods:** Physical behaviours were quantified by integrating the signals from two triaxial accelerometers worn simultaneously during free-living, in a subsample of participants from the Middle-aged Soweto Cohort (MASC) (n=794; mean (SD) age: 53.7 (6.3) years). Acceleration time-series from the two accelerometers were combined and movement-related acceleration derived using Euclidean Norm Minus One (ENMO, in milli-g). This was summarised as total movement volume (mean ENMO) and time spent in non-movement (<28mg), light intensity physical activity (LPA, 28-85 mg) and moderate-vigorous intensity physical activity (MVPA, >85 mg); thigh pitch angle and a sleep diary were used to further divide non-movement time (min/day) spent into sleep, awake sitting/lying, and standing. Socio-demographic factors were self-reported, and weight and height were measured.

**Results:** Mean (SD) wear time for combined thigh-hip accelerometry was 128 (48) hours. Movement volume was 15.0 (6.5) mg for men (n=437; 53.6 (6.2) years) and 12.2 (3.4) mg for women (n=357; 53 (5.8) years). Men spent more time in MVPA and sitting/lying, while women spent more time standing. Age was inversely associated with movement volume, MVPA and LPA in men and women. When compared to their normal weight counterparts, men who were overweight or obese spent less average daily time in MVPA, while women who were overweight or obese spent less time in LPA and more time sitting/lying. Socio-economic status was inversely associated with volume, MVPA and time spent sleeping, and positively associated with time spent sitting/lying, in both men and women.

**Conclusions:** Integrating signals from hip and thigh accelerometers enables characterisation of physical behaviours that can be applied in an African population. Age, female sex, BMI and socio-economic status are inversely associated with physical activity and directly associated with sedentary behaviour.

## Background

It is widely accepted that physical activity is associated with a multitude of health benefits, as reflected in recent public health recommendations encouraging all individuals to move more and sit less (1). Physical inactivity has been identified as a major risk factor for non-communicable diseases (NCDs) (2), the burden of which is nearly equal to the total burden from communicable, maternal, neonatal and nutritional diseases combined in sub-Saharan Africa (3). Increasing urbanisation, life expectancy and population size in sub-Saharan Africa are expected to contribute to this growing burden (4).

One of the challenges in physical activity epidemiology is the measurement of physical behaviour, and there continues to be debate about the use of self-report and objectively measured physical activity (5). It is clear that an understanding of time spent in the various physical activity behaviours is crucial, particularly in low-resourced settings where patterns of physical activity are unique (6). However, the instruments most often used in these settings to measure time spent in physical activity behaviours are questionnaires, which have considerable error.

Accelerometers and other device-based methods provide objective assessment of physical behaviours, avoiding the key issues with self-report, namely recall bias and social desirability bias. Wearable devices also provide more accurate estimates of activity across the whole day, and across the entire intensity spectrum (i.e. from sedentary to vigorous activity), compared to subjective methods which often do not capture light-intensity activity or incidental activity behaviours that are difficult to recall. In particular, hip-worn accelerometers have been suggested as a good method for accurate detection of activity (7), and thigh-worn accelerometers for more detailed postural assessment for classifying sedentary behaviour by discriminating between sitting and standing postures (8). Recent WHO guidelines for physical activity highlight the importance of limiting sedentary behaviour, and increasing activity of any intensity, including light-intensity activity for the prevention of chronic disease and premature mortality (1). Accurately capturing all physical behaviours occurring during the day is hence a priority for public health and physical activity surveillance. Although it is still relatively uncommon for wearable devices to be used in epidemiological studies in Africa (9), there is a need for more precise physical activity data from this region and for global physical activity surveillance in general (10, 11).

Here, we aimed to develop a method of combining the signals from two accelerometers, one worn on the hip and one on the thigh, to quantify physical behaviours as defined by their posture and movement intensity. This allowed us to describe these behaviours by socio-demographic characteristics in a cohort of middle-aged South-African men and women.

## Methods

### Study setting and participants

The Middle-aged Soweto Cohort (MASC) is a longitudinal study of 2031 participants residing in Soweto, Johannesburg, and was designed to identify the determinants of non-communicable disease and type 2 diabetes risk in middle-aged South-African men and women (12). Soweto is an urban township south-west of Johannesburg, with a population of approximately 1.3 million people living in a 200 km^2^ area. The follow-up assessment of this study included objective assessment of physical activity in a subsample of participants which forms the basis of the present cross-sectional analysis. A sample of 502 men and 527 women were invited to participate, and although 99.8% of the sample agreed, accelerometer data were only collected on 839 participants due to the availability of devices. Data were collected between January 2017 and August 2018.

### Anthropometric measures

Height was measured to the nearest 0.1 cm using a stadiometer (Holtain, UK) and weight was measured in light clothing using a digital scale (Dismed, USA) to the nearest 0.1 kg. Body mass index (BMI) was calculated as weight/height^2^ (kg/m^2^).

HIV status was originally acquired at baseline data collection. Participants who were HIV negative at baseline data collection completed an HIV antibody test (Wondfo One Step HIV–1/2, Guangzhou Wondfo Biotech Co., Ltd) during this visit. Participants with a positive HIV result were retained in the study and referred to a clinic.

### Socio-economic status

Socioeconomic status (SES) was quantified using household asset index and education level completed and was assessed using an interviewer-administered questionnaire. Household asset index was calculated by summing assets reported to be owned in the household (out of a total of 12 assets), as has been used in previous studies in this setting (13). Participants reported the highest education level completed, which was categorised into no formal/elementary, secondary or tertiary education levels.

### Physical Activity assessment

Participants were asked to wear two triaxial accelerometers simultaneously; an ActiGraph GT3X+ (ActiGraph, Pensacola, USA) on the hip (ACC_hip_) and an activPAL (PAL Technologies Ltd., Glasgow, UK) on the thigh (ACC_thigh_). At the clinic visit, the ACC_hip_ was fitted on an elastic waistband on the right hip at the midaxillary line and the ACC_thigh_ to the right thigh (anterior midline) using a nitrile sleeve and adhesive dressing (3M Tegaderm). Participants were asked to wear both accelerometers continuously for 7 days and nights, with the exception of the ACC_hip_ being removed for bathing and water-based activities due to the device not being waterproof. Monitors were initialised to record raw triaxial acceleration at 80 Hz and 20 Hz with a dynamic range of +/-6 g and +/-2 g for the ACC_hip_ and ACC_thigh_, respectively. Participants were asked to record their daily sleep (including nap) and wake times using a sleep diary spanning the full measurement period.

### Accelerometer data processing

Following download of the monitors, raw triaxial data were exported from proprietary software (Actilife software, ActiGraph, Pensacola, USA; and activPAL software, PAL Technologies Ltd., Glasgow, UK). Data from both devices was processed using Pampro, an open source software package (14). Data was resampled to 80 Hz and 20 Hz for ACC_hip_ and ACC_thigh_, respectively, and measured acceleration calibrated to local gravity (15). Vector magnitude was calculated from the three axes, and Euclidean Norm Minus One (ENMO) was derived by subtracting 1 g from vector magnitude to remove the gravity component of the acceleration signal and isolate the activity-related signal, expressed in milli-g (mg). Negative values were rounded to zero. High-frequency noise was additionally removed from the ACC_hip_ signal using a 20 Hz low-pass filter; this noise reduction is not possible for ACC_thigh_ due to its lower native sampling frequency.

Periods of non-wear were identified as time segments of low movement variability relative to sensor noise levels, i.e. axis acceleration SD below 13 mg for thigh acceleration (16) and below 4 mg for hip acceleration (17) for ≥1 hour. These periods were removed from the present analysis.

Sample level data was collapsed to 1-minute level data to ensure time synchronisation between the two devices. Furthermore, to account for unexpected time-drifts between the two accelerometers, time segments were set to non-wear if the correlation between hip and thigh acceleration was <0.5.

The time-series of the ACC_thigh_ and ACC_hip_ were combined to derive the average ENMO of the two signals (ENMO_thigh+hip_, main method); the two single-monitor signals were also analysed, making a total of three assessment methods or processing pipelines. One-minute windows were initially classified as either moving or static, based on differing thresholds for ENMO for the three separate pipelines (ACC_thigh+hip_, ACC_thigh_ or ACC_hip_) (Figure 1).

**Figure 1:**
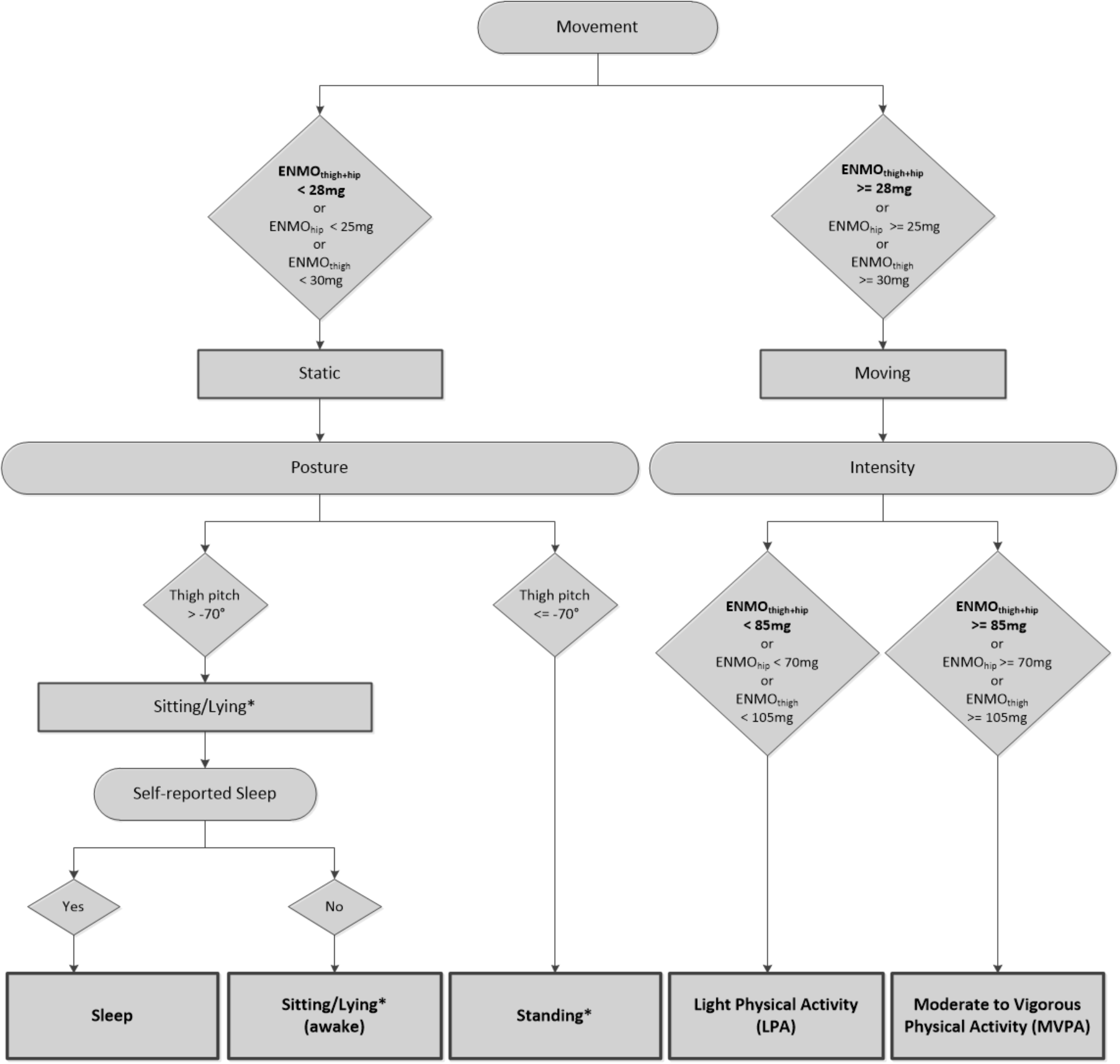
Decision tree for classifying physical behaviour using thigh and hip acceleration signals. *No posture variables generated for the ACC_hip_ signal only.

For time segments classified as moving, magnitude of the acceleration signal(s) (ENMO) was used to classify physical activity intensity into time spent in light physical activity (LPA) and moderate to vigorous physical activity (MVPA). Intensity thresholds were based on ACC_hip_ thresholds (18) and regression within the dataset to derive the equivalent threshold from the ACC_thigh_ signal (from data surrounding the light and moderate threshold regions).

For time classified as static, thigh pitch angle was used for posture-based classification (Figure 1). No specific postures were derived from the ACC_hip_ signal alone due to the poor accuracy of this signal for differentiating lying, sitting and upright postures (19). When the thigh accelerometer is not moving, the measured acceleration is predominantly the gravitational component, from which angles with respect to vertical can be derived. Thigh pitch angle (from -90° to +90°) was calculated (20) along the X axis of ACC_thigh_; a pitch angle of 0° represents the accelerometer being horizontal (perpendicular to gravity), whereas -90° and +90° indicates the accelerometer being vertical (aligned with the gravitational acceleration). When considering anatomical mounting on the thigh, here negative pitch angles indicate an upright position (hip above knee), and hence pitch angle from ACC_thigh_ was used to classify standing and sitting/lying. Adjustment (multiplication by -1) was made to the pitch results where it was evident that the thigh accelerometer had been worn upside-down, i.e. very little time is spent with the knee above the hip, particularly during movement during the day. Due to the horizontal position of the thigh during both sitting and lying, it is difficult to differentiate between these two postures with a thigh-worn monitor.

Finally, to further classify sitting/lying time into sleep time and awake sitting/lying time, the self-reported sleep timings were overlaid onto the accelerometry time-series (Figure 1). Where values were missing for one or more days, either within-person median values for other days were used, or where no sleep data were available for a participant, population sample median times were applied.

All data was plotted and manually/visually verified. An example of the raw signals and derived intensities and postures across a 24-hour period for one participant is displayed in Figure 2.

**Figure 2:**
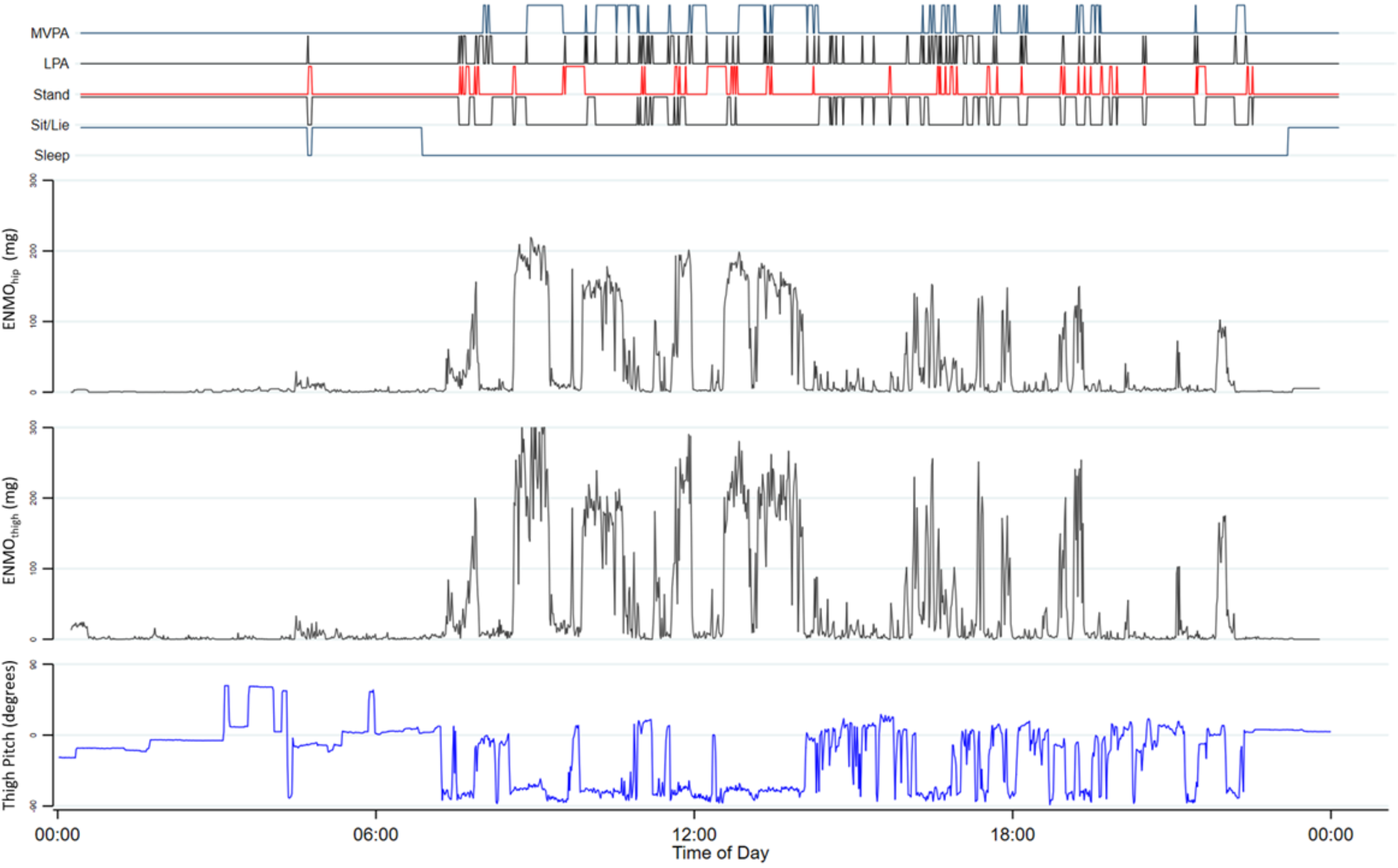
Example of simultaneous hip and thigh acceleration and thigh angles, along with derived intensities and postures across 24-hour period

Data was summarised whilst minimising potential bias caused by imbalance of non-wear (21). Summary results were further consolidated, and the combined (dual) accelerometer signal being used preferentially. Inclusion criteria specified wear time of a minimum of 48 hours with the additional requirement of ≥9 hours cumulative wear in each quadrant (6-hour block) of the day to ensure a balanced representation of the diurnal profile. When these criteria were not met, or wear time from one device was significantly higher than the other, or time synchronisation failed between devices, the signal with the higher wear time was used. Alternatively, ACC_thigh_ was used preferentially in the event that wear time was similar between the signals (<24-hour difference) to allow postural classification.

Final accelerometry outcomes included overall movement volume measures (average ENMO_thigh+hip_ (mg), ENMO_thigh_ (mg) and ENMO_hip_ (mg)), as well as time spent in sleep (min/day), awake sitting/lying (min/day; labelled as sitting/lying from here onwards), standing (min/day), light-intensity physical activity (LPA, min/day) and moderate-vigorous-intensity physical activity (MVPA, min/day). Moving (min/day) is a composite of LPA and MVPA, and static (min/day) is a composite of time spent standing and sitting/lying.

### Statistics

To facilitate direct comparison to other studies using only one accelerometer on either hip or thigh, we derived harmonisation equations by regressing each source acceleration onto the combined acceleration signal from the two anatomical positions, using multi-level time-series regression (level 1; minute-by-minute acceleration, level 2; participant) with linear, square root and square terms for the input signals.

Distributions of person-minutes (time-series level) for key accelerometer signal components are described in histograms by the classified behaviour types across the whole sample.

Descriptive characteristics at participant level are presented as mean (SD) for continuous data and n (%) for categorical data, stratified by sex. Further stratifications included age group (41-49; 50-59; 60-72 years), BMI category according to standard WHO categories (underweight, <18.5 kg/m^2^; normal weight, BMI 18.5-24.9 kg/m^2^; overweight, BMI 25-29.9 kg/m^2^; obesity class I, BMI 30-34.9 kg/m^2^; obesity class II, 35-39.9 kg/m^2^; obesity class III, ≥40.0 kg/m^2^) (22), HIV status (infected, uninfected), and level of education completed (no formal/elementary, secondary or tertiary). Data visualised as box plots represent unadjusted median and interquartile (IQR) ranges, stratified by sex for age categories, BMI categories and SES (education) categories. For household assets, bin scatter plots were used (in 5% bins), adjusted for age.

Sex-stratified multivariable linear regression was used to model the independent associations of age, BMI, HIV status, education level, socio-economic status (asset count) and season of measurement, with physical behaviours.

All statistical analyses were performed using STATA/SE version 16 (StataCorp, TX, USA).

## Results

### Participant characteristics

Complete accelerometry data for both hip and thigh were available on 794 participants (Supplementary Figure 1). A total of 357 women and 437 men were included in the current analysis, with data available from at least one of the two accelerometers. This subsample did not differ significantly from the sample who did not have accelerometry data with respect to BMI and socio-demographic factors, however women included in these analyses were approximately 1.5 years younger than women who were not included (Supplementary Table 1). Descriptive characteristics of the participants in this study are presented in Table 1. Overall, mean (SD) age was 53.7 (6.3) years, with no difference between men and women. Mean (SD) BMI for women was 33.6 (6.9) kg/m^2^ compared to 25.6 (6.0) kg/m^2^ for men, while overweight and obesity were reported in 90% of women and in 51% of men.

**Table 1:** Descriptive Characteristics of the Middle-aged Soweto Cohort (MASC) study (2017-2018).

|  | **Total**<br>**n=794** |  | **Men**<br>**n=437** |  | **Women**<br>**n=357** |  | **p-value** |
| --- | --- | --- | --- | --- | --- | --- | --- |
|  | N or<br>mean | % or<br>SD | N or<br>mean | % or<br>SD | N or<br>mean | % or<br>SD |  |
| **Age (years)** | 53.7 | 6.0 | 53.6 | 6.2 | 53.9 | 5.8 | 0.518 |
| **Age categories:** |  |  |  |  |  |  | 0.179 |
| 41-49 years | 253 | 31.9 | 147 | 33.6 | 106 | 29.7 |  |
| 50-59 years | 387 | 48.7 | 200 | 45.8 | 187 | 52.4 |  |
| 60-72 years | 154 | 19.4 | 90 | 20.6 | 64 | 17.9 |  |
| **BMI (kg/m<sup>2</sup>)** | 29.2 | 7.6 | 25.6 | 6.0 | 33.6 | 6.9 | <0.001 |
| **BMI categories:** |  |  |  |  |  |  | <0.001 |
| <18.5 kg/m<sup>2</sup> | 43 | 5.4 | 41 | 9.4 | 2 | 0.6 |  |
| 18.5-24.9 kg/m<sup>2</sup> | 208 | 26.2 | 175 | 40.0 | 33 | 9.2 |  |
| 25-29.9 kg/m<sup>2</sup> | 204 | 25.7 | 125 | 28.6 | 79 | 22.1 |  |
| 30-34.9 kg/m<sup>2</sup> | 173 | 21.8 | 67 | 15.3 | 106 | 29.7 |  |
| 35-39.9 kg/m<sup>2</sup> | 98 | 12.3 | 18 | 4.1 | 80 | 22.4 |  |
| ≥40 kg/m<sup>2</sup> | 68 | 8.6 | 11 | 2.5 | 57 | 16.0 |  |
| **HIV status:** |  |  |  |  |  |  | 0.617 |
| Infected | 155 | 19.5 | 88 | 20.2 | 67 | 18.8 |  |
| **Education categories:** |  |  |  |  |  |  | 0.001 |
| No formal/Elementary | 74 | 9.3 | 49 | 11.2 | 25 | 7.0 |  |
| Secondary | 595 | 74.9 | 305 | 69.8 | 290 | 81.2 |  |
| Tertiary | 125 | 15.7 | 83 | 19.0 | 42 | 11.8 |  |
| **Socio-economic status:** |  |  |  |  |  |  |  |
| Asset Count | 8.8 | 2.2 | 8.7 | 2.3 | 8.8 | 2.0 | 0.946 |
| **Accelerometry** |  |  |  |  |  |  |  |
| ENMO<sub>thigh+hip</sub> (mg) | 13.8 | 5.5 | 15.0 | 6.5 | 12.2 | 3.4 | <0.001 |
| ENMO<sub>hip</sub> (mg) | 11.5 | 4.2 | 12.5 | 4.8 | 10.3 | 3.0 | <0.001 |
| ENMO<sub>thigh</sub> (mg) | 16.0 | 6.5 | 17.4 | 7.6 | 14.2 | 4.3 | <0.001 |
| Moving (min/day) | 164.9 | 91.1 | 175.7 | 108.9 | 151.7 | 60.4 | <0.001 |
| LPA (min/day) | 118.6 | 78.2 | 118.4 | 95.7 | 118.8 | 49.0 | 0.112 |
| MVPA (min/day) | 46.4 | 36.8 | 57.3 | 42.2 | 32.9 | 22.5 | <0.001 |
| Static (min/day) | 857.6 | 101.6 | 843.8 | 110.0 | 874.5 | 87.4 | <0.001 |
| Standing (min/day) | 223.6 | 98.1 | 195.9 | 86.4 | 257.6 | 101.0 | <0.001 |
| Sitting/Lying (min/day) | 632.9 | 129.4 | 646.9 | 127.3 | 615.8 | 130.0 | <0.001 |
| Sleep (min/day) | 417.5 | 79.9 | 420.5 | 85.3 | 413.8 | 72.8 | 0.281 |

Data are presented as mean (SD) or n(%); p-values are for sex differences.
ENMO = Euclidean Norm Minus One (movement volume); LPA = Light Physical Activity; MVPA = Moderate-to-Vigorous Physical Activity.
Sample sizes where different:
Total: n=777 (ACC<sub>hip</sub>), n = 756 (ACC<sub>thigh</sub>), n =734 (Stand, Sit/Lie), n=778 (sleep diary)
Men: n=428 (ACC<sub>hip</sub>), n = 417 (ACC<sub>thigh</sub>), n =404 (Stand, Sit/Lie), n=433 (sleep diary)
Women: n=349 (ACC<sub>hip</sub>), n = 339 (ACC<sub>thigh</sub>), n =330 (Stand, Sit/Lie), n=345 (sleep diary).

### Movement characteristics at time-series level

Mean (SD) wear time for the hip and thigh accelerometers were 143 (29) and 142 (36) hours, respectively, with the combined signal based on 128 (48) hours of wear. The hip and thigh acceleration signals were highly correlated with each other in the multi-level time-series analyses of 5,493,937 person-minutes of valid double-sensor data (82% explained within-person variance, 77% explained variance between persons), resulting in strong predictions of the combined acceleration signal from either of the two single sources of acceleration, with standard errors of the predictions being 5.4 mg from thigh acceleration and 7.2 mg from hip acceleration (Table 2).

**Table 2:** Relationships (harmonisation equations) between thigh and hip acceleration measures in South African adults.

| Outcome | Prediction equation |
| --- | --- |
| ENMO<sub>thigh+hip</sub> | $1.380617 \times ENMO_{hip} - 0.8988627 \times \sqrt{ENMO_{hip}} - 0.0008704 \times ENMO_{hip}^2$ |
|  | (Within/between person R<sup>2</sup>: 0.94 / 0.92. RMSE = 7.2mg) |
| ENMO<sub>thigh+hip</sub> | $0.8044746 \times ENMO_{thigh} - 0.2011213 \times \sqrt{ENMO_{thigh}} - 0.0000213 \times ENMO_{thigh}^2$ |
|  | (Within/between person R<sup>2</sup>: 0.97 / 0.96. RMSE = 5.4mg) |
| ENMO<sub>hip</sub> | $0.6089491 \times ENMO_{thigh} + 0.4022425 \times \sqrt{ENMO_{thigh}} + 0.0000426 \times ENMO_{thigh}^2$ |
|  | (Within/between person R<sup>2</sup>: 0.82 / 0.77. RMSE =10.8 mg) |

The person-time distribution of hip acceleration, thigh acceleration, and thigh pitch angles are shown by the five classified physical behaviour types in Figure 3. Some aspects of the distributions are defined by the specific decisions in the classification method such as the cut-off for the thigh pitch angle of -70° to separate standing from the other two static postures of sleep and sitting/lying, and therefore by definition, there is no data on the other side of a thigh pitch angle of -70° for sleeping and sitting/lying. Other results reflect the natural unconstrained variation of movement behaviours in this population. For example, participants assume a range of thigh angles around the horizontal position (0°) during sleep and sitting/lying (median (IQR) 2 (−8;9)° and -1 (−14;9)°, respectively), including angles where the knee is almost vertically above the hip. Comparatively, the majority of LPA and MVPA is performed with the knee below the hip (upright position akin to standing) but with LPA displaying a wider range of thigh angles including some approaching horizontal. Similarly, hip and thigh acceleration are by definition restricted to a low range for the three static behaviours but there is still a degree of movement in all three with the thigh displaying a wider range and moving the most compared to the hip. This observation can also be made for LPA and MVPA, which are characterised by more movement at both anatomical locations (note, scale factor 10 larger) compared to the static behaviours. For LPA, median (IQR) hip acceleration is 34 (27;45) mg with most data below 100 mg, and median (IQR) thigh acceleration is 49 (38;66) mg with most data below 120 mg; the theoretical maximal acceleration for both anatomical locations is 170 mg, ie twice the combined intensity cutpoint. For MVPA, median (IQR) hip acceleration is 107 (84;139) mg with not much time above 300 mg, whereas the thigh typically moves more during MVPA, median (IQR) acceleration at 161 (127;208) mg and most data below 400 mg.

**Figure 3:**
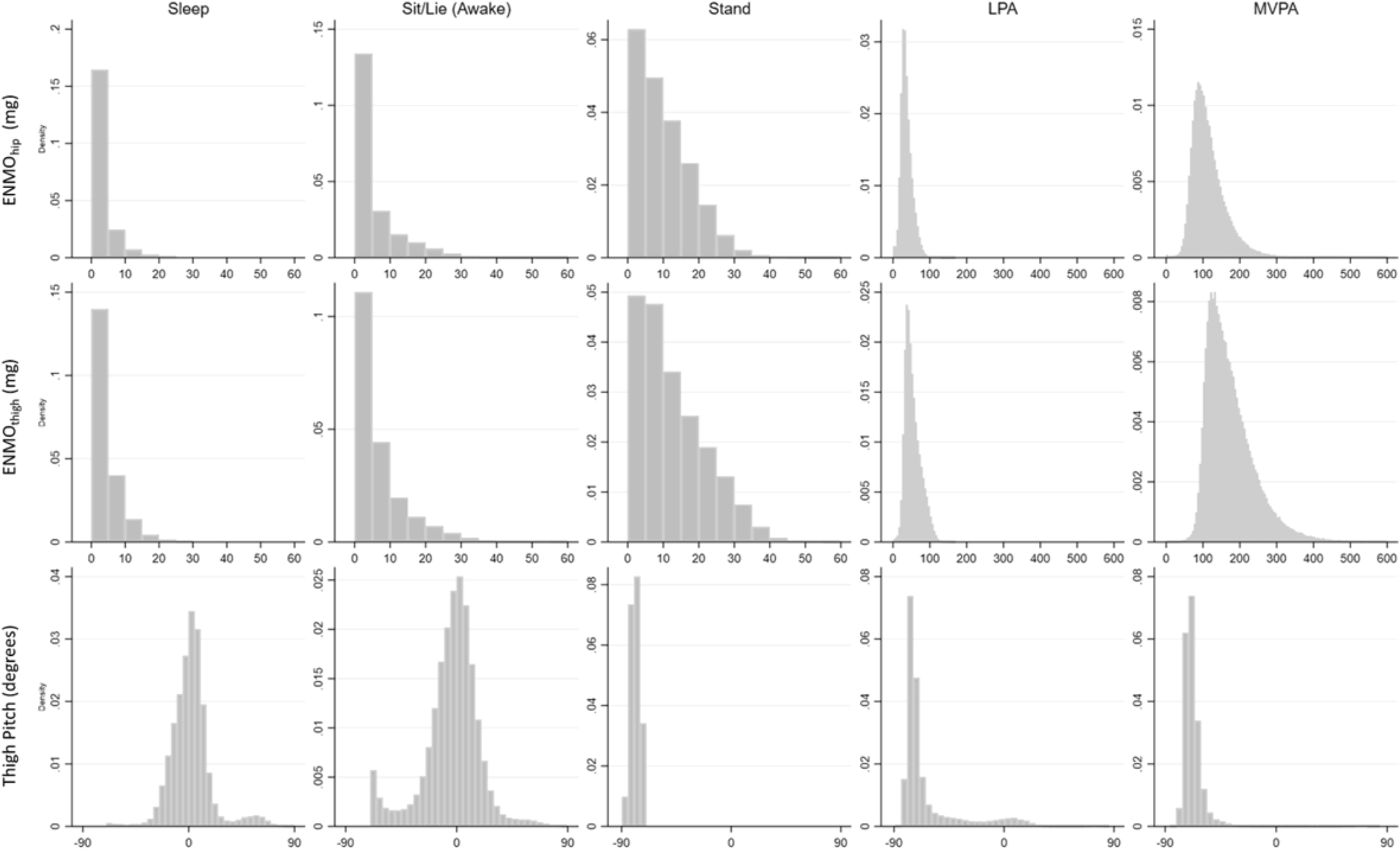
Person-time distribution of hip acceleration, thigh acceleration and thigh angles across classified behaviour types. ENMO plots only display values up to 600mg.

### Descriptive epidemiology of physical behaviours at participant level

The descriptive characteristics, including the physical behaviours, of the total sample and men and women separately are presented in Table 1. Physical behaviours were different between the sexes. Men spent more time moving than women, particularly at higher intensity, as can be seen from higher values of ENMO and more time spent in MVPA. Women spent more time standing and less time sitting/lying compared to men. There were no sex differences for time spent sleeping or in LPA.

The box plots (unadjusted) of all the physical behaviours by three age categories and stratified by sex are presented in Figure 4. For both men and women, ENMO, LPA and MVPA decreased with age, with no associations for time spent standing, sitting/lying, and sleeping. In the multivariable analysis with adjustment for BMI, HIV status, SES and season of measurement, ENMO and time spent in MVPA was lower in the oldest age group (60-72 years) compared to the youngest age group (40-49 years) in both men (Table 3a) and women (Table 3b). Both men and women in the oldest age group spent ∼13 min/day less in MVPA compared to the youngest age group, but differences in time spent in LPA and sitting/lying were only shown in women where the oldest age group spent 19 min/day less in LPA and 46 min/day more time in sitting/lying compared to the youngest age group. Men between the ages of 50 and 59 years slept an average of 22 minutes more per day than men between the ages of 40 and 49 years. Similar associations with age were observed when only considering hip acceleration (Supplementary Table 2). While HIV status was not associated with any of the physical behaviours in men, women living with HIV spent 30min/day less in standing that women not infected by HIV (Table 3b).

**Figure 4:**
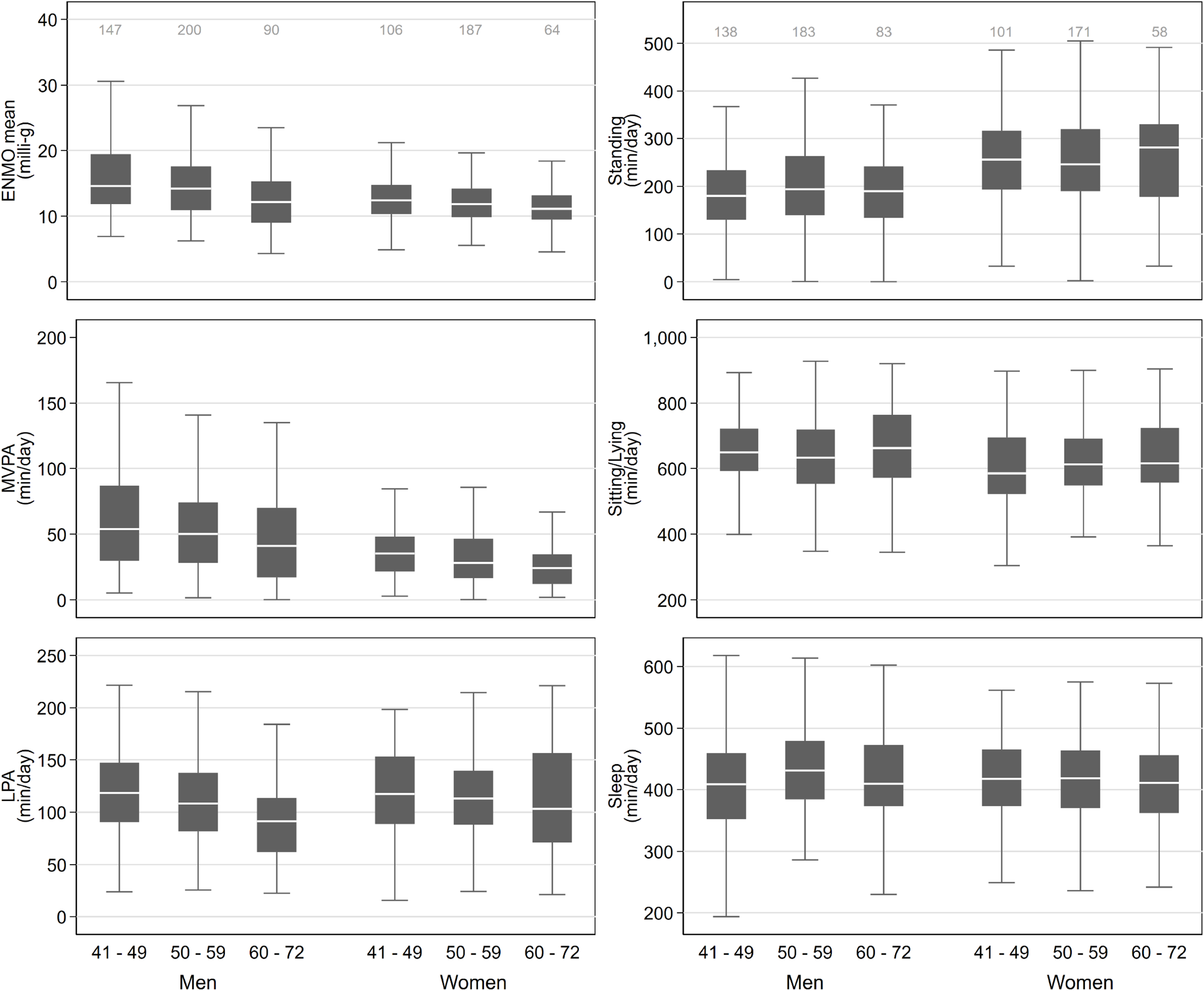
Physical activity behaviours by age and sex in the Middle-aged Soweto Cohort (MASC) study 2017-2018. Sample sizes for ENMO, MVPA, LPA, and Sleep are the same and only displayed in the ENMO panel; the sample sizes for Standing and Sitting/Lying are also the same and displayed in the Standing panel.

**Table 3a:** Multivariable regression analysis of physical behaviours in men

|  | **ENMO (mg)** | **MVPA (min/day)** | **Light PA**<br>**(min/day)** | **Standing**<br>**(min/day)** | **Sitting/Lying**<br>**(min/day)** | **Sleep**<br>**(min/day)** |
| --- | --- | --- | --- | --- | --- | --- |
|  | Beta (95% CI) | Beta (95% CI) | Beta (95% CI) | Beta (95% CI) | Beta (95% CI) | Beta (95% CI) |
| **Age (years)** |  |  |  |  |  |  |
| 40-49 | Reference | Reference | Reference | Reference | Reference | Reference |
| 50-59 | -1 (-2.2; 0.3) | -5.9 (-14.2; 2.4) | -0.9 (-21.6; 19.7) | 7.6 (-11.9; 27.1) | -23.5 (-51.6; 4.5) | 21.5 (3.5; 39.5) |
| 60-72 | -3.0** (-4.6; -1.4) | -12.6* (-23.1; -2.2) | -14.3 (-40.3; 11.6) | -11.5 (-36.1; 13.1) | 12.3 (-23.2; 47.7) | 18.7 (-3.8; 41.3) |
| **BMI (kg/m<sup>2</sup>)** |  |  |  |  |  |  |
| <18.5 | -2.5* (-4.5; -0.5) | -6 (-19.2; 7.2) | -15.9 (-48.8; 17) | -25.9 (-57.2; 5.3) | 4.8 (-40.4; 49.9) | 38.0** (9.4; 66.6) |
| 18.5-24.9 | Reference | Reference | Reference | Reference | Reference | Reference |
| 25-29.9 | -1.9** (-3.3; -0.6) | -16.2** (-25.1; -7.3) | 6.3 (-15.8; 28.5) | 18.9 (-2.1; 39.9) | 7.5 (-22.9; 37.8) | -17.2 (-36.4; 2.1) |
| 30-34.9 | -3.0** (-4.7; -1.3) | -21.4** (-32.5; 10.3) | -15.0 (-42.6; 12.7) | 5.5 (-20.8; 31.8) | 58.4** (20.5; 96.4) | -30.2* (-54.2; -6.1) |
| 35-39.9 | -1.8 (-4.8; 1.1) | -6.6 (-25.4; 12.1) | -18.2 (-64.7; 28.4) | -29.7 (-74.6; 15.1) | 50.4 (-14.3; 115.1) | 0.2 (-40.2; 40.7) |
| >=40 | -0.9 (-4.5; 2.7) | -29.8* (-53.3; -6.3) | 123.5** (65; 181.9) | -53.2 (-106.7; 0.3) | 6.2 (-71; 83.3) | -47.1 (-97.9; 3.7) |
| **HIV status** |  |  |  |  |  |  |
| HIV uninfected | Reference | Reference | Reference | Reference | Reference | Reference |
| HIV infected | -1.1 (-2.5; 0.3) | -4.5 (-13.8; 4.9) | -16.5 (-39.7; 6.7) | -7.9 (-29.8; 13.9) | 14.1 (-17.5; 45.7) | 15.8 (-4.3; 36) |
| **Education level** |  |  |  |  |  |  |
| No formal/elementary | Reference | Reference | Reference | Reference | Reference | Reference |
| Secondary | 1.6 (-0.2; 3.4) | 10.4 (-1.3; 22) | 12.3 (-16.5; 41.2) | 7.1 (-20.1; 34.4) | -14.1 (-53.4; 25.2) | -10.2 (-35.3; 14.9) |
| Tertiary | 1.1 (-1.1; 3.2) | 8.3 (-5.8; 22.4) | 21.3 (-13.8; 56.4) | -5.4 (-38.3; 27.4) | 1 (-46.4; 48.3) | -16.8 (-47.3; 13.7) |
| **Socio-economic status** |  |  |  |  |  |  |
| Asset count in household<sup>†</sup> | -0.5** (-0.7; -0.2) | -6.2 ** (-7.9; -4.6) | 1.5 (-2.5; 5.6) | -1 (-4.9; 2.8) | 9.1** (3.5; 14.7) | -3.4 (-7.0; 0.1) |
| **Seasonality** |  |  |  |  |  |  |
| Spring | 0.3 (-0.6; 1.2) | 4.2 (-1.5; 9.9) | 3.4 (-10.8; 17.5) | -10.5 (-24.1; 3.1) | 13.3 (-6.4; 32.9) | -13.0* (-25.3; -0.7) |
| Winter | 0.2 (-0.7; 1.1) | 2.9 (-2.8; 8.5) | -3.4 (-17.5; 10.7) | 6.2 (-7.3; 19.8) | -19.6* (-39.1; 0) | 15.5* (3.3; 27.8) |
| **Constant<sup>†</sup>** | 16.4** (14.3; 18.5) | 63.0** (49.3; 76.6) | 114.2** (80.2;148.1) | 186.2** (154.3; 218.2) | 659** (613.7; 705.9) | 412.4** (382.9; 441.9) |

ENMO, MVPA and LPA estimates are based on ENMO<sub>thigh+hip</sub> time-series where available, and (scaled) from either ENMO<sub>thigh</sub> or ENMO<sub>hip</sub> where not.
\*\* p<0.01, \* p<0.05. <sup>†</sup>Centred at 9 assets. Seasonal parameters are continuous cosinor coefficients, oscillating from -1 to +1, peaking at October 1st (Spring) and July 1st (Winter).

**Table 3b:** Multivariable regression analysis of physical behaviours in women

|  | **ENMO (mg)** | **MVPA (min/day)** | **Light PA**<br>**(min/day)** | **Standing**<br>**(min/day)** | **Sitting/Lying**<br>**(min/day)** | **Sleep**<br>**(min/day)** |
| --- | --- | --- | --- | --- | --- | --- |
|  | Beta (95% CI) | Beta (95% CI) | Beta (95% CI) | Beta (95% CI) | Beta (95% CI) | Beta (95% CI) |
| **Age (years)** |  |  |  |  |  |  |
| 40-49 | Reference | Reference | Reference | Reference | Reference | Reference |
| 50-59 | -0.4 (-1.4; 0.5) | -3.7 (-9.1; 1.8) | -6.1 (-18.5; 6.4) | -8.7 (-34.4; 17) | 24.9 (-7.3; 57.1) | -8.3 (-26.8; 10.2) |
| 60-72 | -2.0** (-3.3; -0.6) | -13.0** (-20.4; -5.5) | -18.6* (-35.5; -1.7) | -3.3 (-38.6; 32) | 46.0 (1.7; 90.3) | -15.4 (-40.6; 9.7) |
| **BMI (kg/m<sup>2</sup>)** |  |  |  |  |  |  |
| <18.5 | -4.7 (-9.9; 0.5) | -24.6 (-55.2; 6.1) | -72.7* (-142.2; -3.1) | 19 (-120.6; 158.6) | 41.7 (-133.3; 216.8) | 40.2 (-63.3; 143.7) |
| 18.5-24.9 | Reference | Reference | Reference | Reference | Reference | Reference |
| 25-29.9 | -0.7 (-2.2; 0.8) | 0.3 (-8.3; 9.0) | -27.6** (-47.3; -7.8) | -12.1 (-52.0; 27.8) | 55.0* (5.0; 104.9) | -19.5 (-48.8; 9.9) |
| 30-34.9 | -0.6 (-2.1; 0.8) | 0.6 (-7.7; 9.0) | -13.1 (-31.9; 5.8) | -13.0 (-51.6; 25.5) | 44.0 (-4.4; 92.3) | -19.0 (-47.1; 9.1) |
| 35-39.9 | -2.0** (-3.5; -0.5) | -6.9 (-15.6; 1.7) | -27.8** (-47.4; -8.3) | -62.8** (-102.3; -23.2) | 111.3** (61.8; 160.8) | -13.8 (-42.9; 15.3) |
| >=40 | -1.9* (-3.5; -0.3) | -8.9 (-18.0; 0.3) | -34.8** (-55.6; -14.1) | -95.4** (-137.9; -52.9) | 146.7** (93.4; 199.9) | -9.3 (-40.2; 21.6) |
| **HIV status** |  |  |  |  |  |  |
| HIV uninfected | Reference | Reference | Reference | Reference | Reference | Reference |
| HIV infected | -0.6 (-1.7; 0.5) | -0.5 (-6.7; 5.7) | -7.4 (-21.4; 6.6) | -30.3* (-58.9; -1.7) | 21.3 (-14.5; 57.2) | 13.4 (-7.5; 34.3) |
| **Education level** |  |  |  |  |  |  |
| No formal/elementary | Reference | Reference | Reference | Reference | Reference | Reference |
| Secondary | -0.2 (-1.8; 1.4) | -1.0 (-10.2; 8.2) | 10.9 (-10; 31.8) | 13.1 (-29.9; 56.2) | -5.7 (-59.7; 48.4) | -21.0 (-52.1; 10.1) |
| Tertiary | -1.6 (-3.6; 0.4) | -9.1 (-20.5; 2.3) | -0.6 (-26.4; 25.3) | -3.1 (-57.3; 51.0) | 60.9 (-6.9; 128.8) | -46.2* (-84.6; -7.7) |
| **Socio-economic status** |  |  |  |  |  |  |
| Asset count in household<sup>†</sup> | -0.2* (-0.4; 0) | -2.9** (-4.1; -1.8) | 1.2 (-1.4; 3.8) | -2.9 (-8.3; 2.5) | 9.0** (2.2; 15.8) | -3.6 (-7.4; 0.3) |
| **Seasonality** |  |  |  |  |  |  |
| Spring | -0.1 (-0.7; 0.4) | 0.1 (-2.9; 3.1) | -0.6 (-7.4; 6.3) | 7.2 (-7.1; 21.5) | 4.9 (-13.0; 22.8) | -12.0* (-22.2; -1.8) |
| Winter | -0.6 (-1.3; 0) | -2.7 (-6.3; 1.0) | -8.4* (-16.6; -0.2) | 10.7 (-6.4; 27.7) | -6.5 (-27.8; 14.8) | 4.1 (-8.1; 16.3) |
| **Constant<sup>†</sup>** | 14.6** (12.4; 16.9) | 41.7** (28.8; 54.7) | 141.3** (111.8; 170.7) | 292.0 (231.6; 352.4) | 516.9** (441.1; 592.6) | 454.8** (411.0; 498.6) |

ENMO, MVPA and LPA estimates are based on ENMO<sub>thigh+hip</sub> time-series where available, and (scaled) from either ENMO<sub>thigh</sub> or ENMO<sub>hip</sub> where not.
\*\* p<0.01, \* p<0.05. <sup>†</sup> Centred at 9 assets. Seasonal parameters are continuous cosinor coefficients, oscillating from -1 to +1, peaking at October 1st (Spring) and July 1st (Winter).

Figure 5 presents box-plots of the physical behaviours stratified by BMI category. They show lower levels of MVPA in the higher BMI categories in men, and similarly for LPA in the women. No clear pattern with BMI was seen for standing in men, whereas women in higher BMI categories stood less than women in lower BMI categories. Time spent sitting was higher at higher BMI levels in both men and women, and underweight and normal weight individuals tended to sleep more. Multivariable analysis shows that when compared to the normal weight category, men who were overweight or obese had significantly lower ENMO (1.9 and 3 mg) and spent less average daily time in MVPA (16 and 21 min) and sleeping (17 and 30 min), and more time standing (18.9 min in the men who were overweight) (Table 3a). When compared to normal weight women, women who were overweight or obese spent significantly less time in LPA (27-35 mins/day) and standing, and more time sitting/lying. When only considering hip acceleration, similar patterns were observed in men and women (Supplementary Table 2).

**Figure 5:**
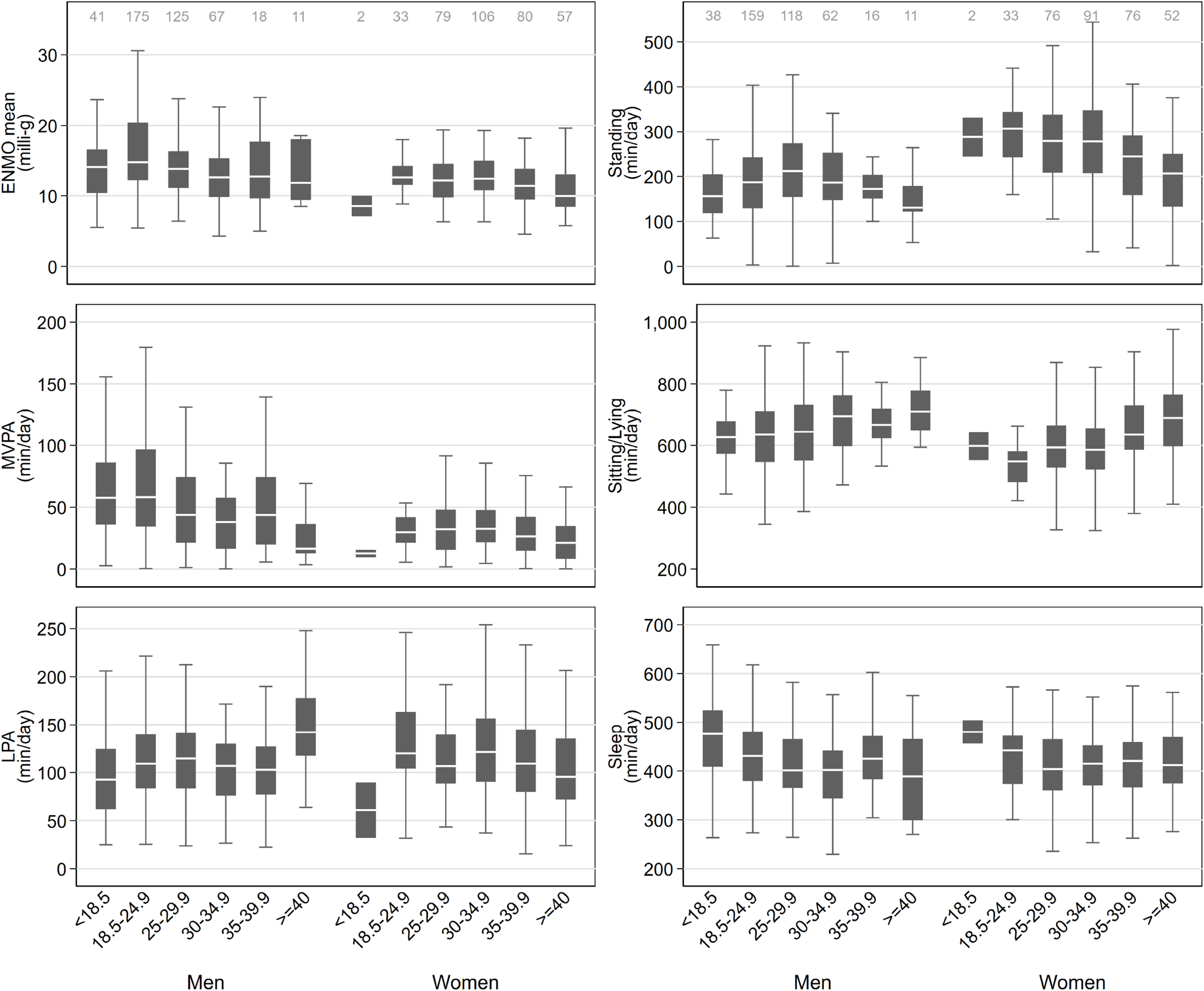
Physical activity behaviours by BMI (kg/m^2^) and sex. The Middle-aged Soweto Cohort (MASC) study 2017-2018. Sample sizes for ENMO, MVPA, LPA, and Sleep are the same and only displayed in the ENMO panel; the sample sizes for Standing and Sitting/Lying are also the same and displayed in the Standing panel.

The variation in PA behaviours by three education categories are presented as box plots in Figure 6. The patterns of associations observed for men were lower PA (ENMO, MVPA and LPA) and more time sleeping in those with no formal education compared to those more educated. Women with higher educational attainment spent less time in MVPA, less time sleeping, and more time sitting compared to those less educated. Multivariable analysis shows that when compared to no formal/elementary education, men with secondary education were more active overall (ENMO difference of 1.6 mg) and also spent more time in MVPA (10 min/day). Women with a tertiary education spent less time sleeping (46 min) compared to women with no formal/elementary education. Similar associations with both ENMO and MVPA were observed in the men and women are observed when only considering hip acceleration (Supplementary Table 2).

**Figure 6:**
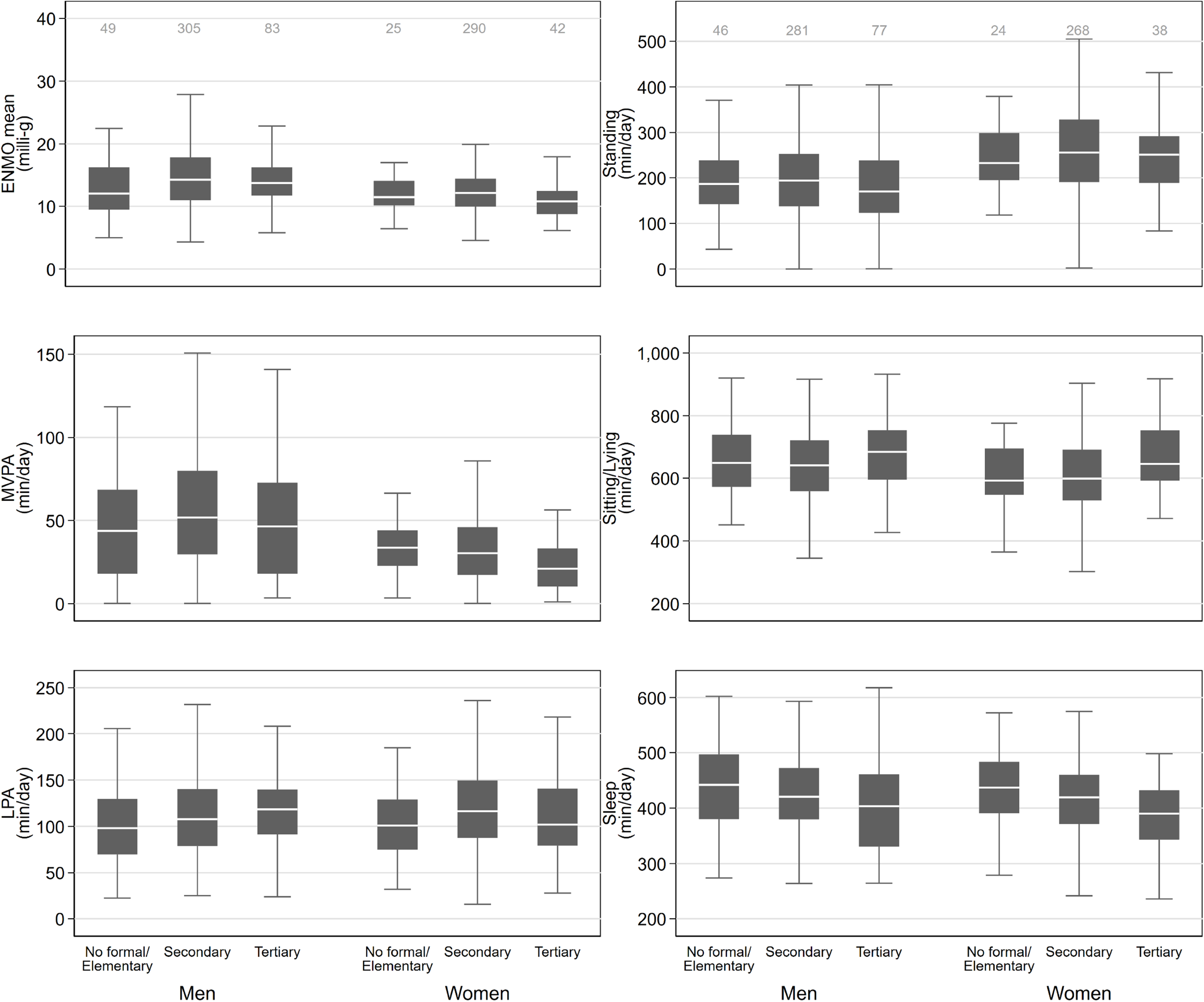
Physical activity behaviours by education and sex. The Middle-aged Soweto Cohort (MASC) study 2017-2018. Sample sizes for ENMO, MVPA, LPA, and Sleep are the same and only displayed in the ENMO panel; the sample sizes for Standing and Sitting/Lying are also the same and displayed in the Standing panel.

The number of assets, a measure of SES, and its association with physical behaviours in men and women are presented in bin scatter diagrams in Figure 7. These show a similar trend in both men and women, with a higher asset count associated with lower MVPA and sleep, and more time sitting/lying. Multivariate analyses show similar associations in both men and women, with a higher asset count associated with lower ENMO (0.5 and 0.2 mg/asset), less daily time in MVPA (6.2 and 2.9 min/asset) and sleeping (3.4 and 3.6 min/asset), and more time spent sitting/lying (9.1 and 9.0 min/asset). Similar associations were observed in both men and women when only considering hip acceleration (Supplementary Table 2).

**Figure 7:**
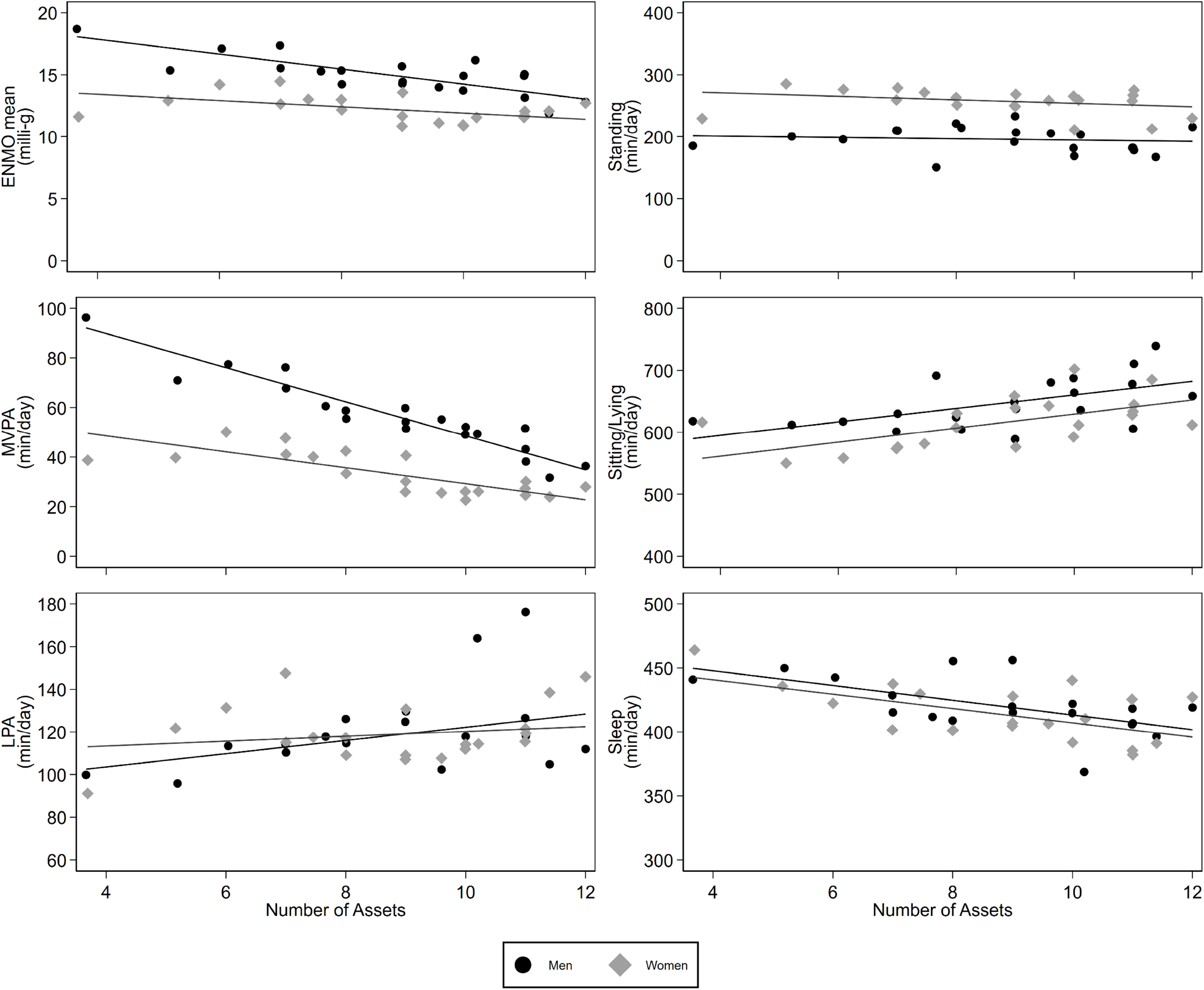
Association between household assets and physical activity behaviours, stratified by sex. The Middle-aged Soweto Cohort (MASC) study 2017-2018

## Discussion

In this study we have developed a method for combining the signals from hip and thigh mounted accelerometers to describe physical behaviours in a middle-aged cohort of men and women living in urban South Africa. This simple method is based on first-principles which discriminates sitting from standing postures using the thigh pitch angle, and a combination of hip and thigh acceleration quantifies different intensities of activity. Using this method, we have shown that men spend nearly double the amount of time in daily MVPA compared to women, but also more time in awake sitting or lying. In identifying factors associated with different physical behaviours we have shown clear associations with age, BMI and socio-economic status, with some of these associations different in men and women.

Integrating accelerometry data from multiple anatomical locations, such as the hip and thigh, has advantages in terms of combining methodological strengths from both methods, to optimise accuracy of derived physical activity subcomponents (7). Previous research has used multiple accelerometry sensors placed at different anatomical locations, from which information is wired into, and processed by, one centralised unit (23-26). Due to this complexity, utility of such devices for long-term monitoring of physical behaviours during free-living in large-scale studies is limited, and often proprietary algorithms are used for activity type classification (27). On the contrary, modern research-grade accelerometers allow for storage of raw data over prolonged monitoring times. Raw acceleration data, when calibrated to local gravity and expressed in mg, are universal and hence device independent, and even derived angles relative to the gravitational acceleration gradient (ie, vertical) during periods of little movement can be considered universal. Compared to device-specific proprietary algorithms, open-source algorithms applied to raw data allow more flexibility for researchers for optimisation and are also applicable across different devices. Algorithmic integration of raw data from such accelerometers collected from different anatomical positions therefore opens up new possibilities to optimise measurement of habitual physical activity behaviour types in large scale studies. Thigh pitch, as derived from thigh accelerometry, has shown great utility for accurate discrimination of sitting/lying postures from upright postures (28), and outperforms hip accelerometry for this classification task, using direct observation as the criterion (29, 30). On the contrary, hip accelerometry appears to be better for the assessment of whole-body acceleration and hence movement intensity, likely given the closer proximity to the body`s centre of mass. However, as we show in the present study, the acceleration from both these anatomical locations are highly correlated. Some studies have combined data from separate hip- and thigh-mounted accelerometers in an algorithm-based manner for discrimination between specific activity types, such as walking, running, cycling, walking stairs, sitting, lying and standing (27, 31). So far, these have mainly been applied in Scandinavian working populations (32, 33). In African settings, objective assessment of activity is limited (9), and non-existent using multi-accelerometer methods. Developing a dual-accelerometer methodology to quantify physical behaviours in an under-studied population and describing the socio-demographic factors associated with these behaviours contributes to the limited literature available in African populations.

As with epidemiological studies in other populations including South Africa, we have shown that men are more physically active than women of the same age (10, 34-39). Although this difference was observed for total movement volume as well as MVPA, there was no difference in time spent in LPA between the sexes. In African countries, occupational and ambulatory activity make the largest contribution to overall physical activity, with leisure time activity contributing very little (6, 40, 41). Ambulatory activity, a requirement of both men and women as a means of transport in a setting such as Soweto, is typically performed at a light intensity, however occupational and leisure-time activity, normally performed at a higher intensity, is more likely to be undertaken by men in these low-resourced settings (6).

A 2016 systematic review of objectively and subjectively measured sedentary time from studies conducted in North America, Europe, Australia, New Zealand and Asia, reported that in the studies that examined sex differences in sedentary time, most reported that women were more sedentary than men (42). Conversely, data from the Caribbean, a region predominantly made up of developing countries, reported no sex difference in sedentary time (37). They defined sedentary time as time spent at < 1.5 MET which may include standing time and reported this to be ∼8 hours/day, which was lower than the current study which reported 10-11 hours/day of sitting/lying, and a further 3-4 hours of standing time. Combining signals from a hip-mounted accelerometer and a thigh-mounted accelerometer which provides additional information on thigh pitch angle, we were able to not only capture body movement more comprehensively but also differentiate sitting/lying from standing and determine the socio-demographic factors associated with a range of physical behaviours including those static postures. Women in our study stand for an average of just over 4 hours/day which is significantly longer than men who stand for about 3 hours/day, while time spent in awake sitting/lying was higher in men than women, although both were over 10 hours a day. Associations between self-reported sitting and standing time, and all-cause mortality, have been explored suggesting every additional hour of sitting time above 8 hours/day is associated with a 4% higher mortality risk, and standing for more than 8 hours/day is associated with a 24% reduced mortality risk compared to standing 2 hours/day (43, 44). Chastin et al., (45) have highlighted the limitations of self-report measures of sedentary time, an illustration of which may be the mismatch between the median sitting time of 3 hours a day reported by Gradidge et al., (40) in the same cohort of women from Soweto using self-report, and the objectively assessed sitting time reported in the current study, albeit 12 years later. We have also shown that while BMI is associated with time spent standing as well as time spent sitting/lying in men and women, age and SES were only associated with time spent sitting/lying. This confirms the importance of discriminating between these two behaviours in this population.

Although increasing physical activity and reducing sedentary time are promoted as methods of reducing body weight and the risk of overweight and obesity, this relationship is likely to be bi-directional (46-48). In the current study, men who were overweight or obese (BMI 25-35 kg/m^2^) had lower total volume of physical activity and spent less time in MVPA compared to their normal weight counterparts. In women, higher BMI was more closely associated with less time spent in LPA and more time spent sitting/lying. This may just reflect differences in physical behaviour patterns between the sexes in this population. When compared to men with the same BMI (>35 kg/m^2^), women spent more than double the amount of time sitting/lying, and when these women were compared to their normal weight counterparts, they spent 1-1.5 hours less time standing and 2-2.5 hours more sitting/lying.

Various South African studies in adolescents and adults have confirmed that socio-economic status is closely linked to physical activity patterns (38, 40, 49-51). Findings from the current study report that a higher asset count, a well-recognised measure of socio-economic status, was associated with a lower total volume of physical activity, less time in MVPA and more time sitting/lying, in both men and women. The positive association between SES and sedentary time has been confirmed by other South African studies including a study of adolescents from rural South Africa (49, 50), however the adolescent study also reported that lower SES was associated with lower MVPA, contrary to the findings of the current study. The discrepancy in these findings may be due to the use of self-reported physical activity in the adolescent study as well as SES being measured at the maternal, household and community level rather than the individual level. Similarly, in a Caribbean sample between 25 and 54 years of age, education level and occupational grade was inversely associated with objectively measured physical activity and positively associated with sedentary time (37). What is apparent is that the relationship between SES and physical behaviours is complex as physical activity patterns and volumes differ between urban and rural populations as well as between low-middle and high-income countries.

Irrespective of country or setting, it is well accepted that age is a significant correlate of physical activity. In the current study this appears to be more marked for both volume (ENMO) and higher intensity physical activity (MVPA) in older men and women (between 60 and 72 years of age). Findings from the HAALSI study in rural South African adults over the age of 40 years identified age as a significant correlate of self-reported physical activity with adults over the age of 70 years to be less likely to meet physical activity guidelines than those between 40 and 49 years of age (52).

Although there is an increasing interest in the association between sleep duration and quality, and disease risk in South African women, to our knowledge this is the first study reporting sleep duration in South African men. Our measure of sleep was a combination of self-report via the sleep diary and the accelerometry data indicating absence of movement and a sitting/lying posture, with the mean sleep duration being about 7 hours/day for both men and women. This is approximately one hour less than self-reported data from a cohort of younger black women from Cape Town, in which sleep was associated with weight status, with women sleeping less than 7 hours/day being less likely to present with obesity than those sleeping 7-9 hours (53). In the current study BMI was not associated with sleep in the women, however when compared to normal weight men underweight men spent more time sleeping, while men who were overweight or obese spent less time spent sleeping than their normal weight counterparts.

A major strength of the study includes the use of objective methods to measure physical behaviours, and particularly integrating and combining raw sensor data from accelerometers worn on different anatomical locations. Similar associations to other epidemiological studies confirm the robustness of this approach and provides further detail on postural differentiation of sedentary time. Using this integrated methodology, we were able to explore individual and household level correlates of physical behaviours which has previously only been described in women using either self-report or single accelerometer methods, however due to the cross-sectional nature of the study the direction of associations with modifiable determinants cannot be confirmed. Further, although objective methods are preferable when measuring physical activity to avoid issues such as recall bias, these methods are not able to distinguish the domains in which physical behaviours are performed such as leisure time and occupation. Physical activity time spent in these domains is important particularly in settings where physical activity patterns may differ to high income settings. Domain-specific information could help in designing effective interventions in these settings. Another limitation of the current study is the use of sleep diaries to determine awake and sleep time. Sleep diaries are also prone to recall bias as well as reporting time in bed rather than sleep duration, however the combination of sleep diaries and accelerometry may have reduced misclassification errors.

## Conclusions

In this study we have described a method of integrating signals from hip and thigh accelerometers to more comprehensively describe physical behaviours of middle-aged men and women from urban South Africa. Identifying factors associated with physical activity and sedentary behaviour contributes to our understanding of lifestyle behaviours in this population and provides possible areas for intervention.

## Data Availability

All data produced in the present study are available upon reasonable request to the authors

## Abbreviations

MASC: Middle-aged Soweto Cohort
ENMO: Euclidean Norm Minus One
LPA: light intensity physical activity
MVPA: moderate-vigorous intensity physical activity
BMI: Body mass index
NCDs: non-communicable diseases
SES: Socioeconomic status
ACC_hip_: hip accelerometer
ACC_thigh_: thigh accelerometer
ENMO_thigh+hip_: average ENMO from hip and thigh accelerometers
HAALSI: Health and Aging in Africa study

## Ethics approval and consent to participate

The study was conducted in accordance with the tenets of the Helsinki declaration and was approved by the Human Research Ethics Committee (HREC) (Medical) of the University of the Witwatersrand (clearance certificate NO. M160604 and M160975). Prior to inclusion in the study all the procedures and possible risks associated with the study were explained and all participants signed a consent form. All the tests and procedures were carried out at the South African Medical Research Council/Wits Developmental Pathways for Health Research Unit at the Chris Hani Baragwanath Hospital in Soweto, Johannesburg, South Africa.

## Consent for publication

Not applicable.

## Availability of data and materials

The datasets used and/or analyses during the current study are available from the corresponding author on reasonable request.

## Authors contributions

LKM, JHG and SB conceived the study, and KWe, AS and SB processed and analysed the data. All authors drafted and revised the manuscript and read and approved the final version.

## Acknowledgements

We are grateful to all MASC participants as well as DPHRU field staff.

## Additional files

**Additional file 1 – Figure S1** (.tiff)

**Figure S1 title:** Participant/Data flow

**Additional file 2 – Table S1** (.docx)

**Table S1 title:** Descriptive characteristics of sub-sample included and excluded from the current study

**Additional file 3 – Table S2** (.docx)

**Table S2 title:** Multivariable analysis of physical activity volume and time in MVPA using hip-worn accelerometer signal (ACC_hip_) only

## References

1. Bull FC, Al-Ansari SS, Biddle S, Borodulin K, Buman MP, Cardon G, et al. World Health Organization 2020 guidelines on physical activity and sedentary behaviour. Br J Sports Med. 2020;54(24):1451–62.

2. Lee IM, Shiroma EJ, Lobelo F, Puska P, Blair SN, Katzmarzyk PT. Effect of physical inactivity on major non-communicable diseases worldwide: an analysis of burden of disease and life expectancy. The Lancet. 2012;380(9838):219–29.

3. Gouda HN, Charlson F, Sorsdahl K, Ahmadzada S, Ferrari AJ, Erskine H, et al. Burden of non-communicable diseases in sub-Saharan Africa, 1990–2017: results from the Global Burden of Disease Study 2017. The Lancet Global Health. 2019;7(10):e1375–e87.

4. Gyasi RM, Phillips DR. Aging and the rising burden of noncommunicable diseases in sub-Saharan Africa and other Low-and Middle-Income Countries: A Call for holistic action. The Gerontologist. 2020;60(5):806–11.

5. Dowd KP, Szeklicki R, Minetto MA, Murphy MH, Polito A, Ghigo E, et al. A systematic literature review of reviews on techniques for physical activity measurement in adults: a DEDIPAC study. International Journal of Behavioral Nutrition and Physical Activity. 2018;15(1):1–33.

6. Guthold R, Louazani SA, Riley LM, Cowan MJ, Bovet P, Damasceno A, et al. Physical activity in 22 African countries: results from the World Health Organization STEPwise approach to chronic disease risk factor surveillance. Am J Prev Med. 2011;41(1):52–60.

7. Cleland I, Kikhia B, Nugent C, Boytsov A, Hallberg J, Synnes K, et al. Optimal placement of accelerometers for the detection of everyday activities. Sensors. 2013;13(7):9183–200.

8. Tremblay MS, Aubert S, Barnes JD, Saunders TJ, Carson V, Latimer-Cheung AE, et al. Sedentary behavior research network (SBRN)–terminology consensus project process and outcome. International journal of behavioral nutrition and physical activity. 2017;14(1):1–17.

9. Brage S, Assah F, Msyamboza KP. Quantifying population levels of physical activity in Africa using wearable sensors: implications for global physical activity surveillance. BMJ Open Sport & Exercise Medicine. 2020;6(1):e000941.

10. Hallal PC, Andersen LB, Bull FC, Guthold R, Haskell W, Ekelund U. Global physical activity levels: surveillance progress, pitfalls, and prospects. The Lancet. 2012;380(9838):247–57.

11. Westgate K, Ridgway C, Rennie K, Strain T, Wijndaele K, Brage S. Feasibility of incorporating objective measures of physical activity in the STEPS program. A pilot study in Malawi. Geneva: World Health organization; 2019.

12. Goedecke JH, Nguyen K, Kufe C, Masemola M, Chikowore T, Mendham AE, et al. Waist circumference thresholds predicting incident dysglycemia and type 2 diabetes in Black African men and women. Preprint. medrxiv. 2021. DOI: 10.1101/2021.10.18.21265125

13. Munthali RJ, Manyema M, Said-Mohamed R, Kagura J, Tollman S, Kahn K, et al. Body composition and physical activity as mediators in the relationship between socioeconomic status and blood pressure in young South African women: a structural equation model analysis. BMJ Open. 2018;8(12):e023404.

14. Medical Research Council. Pampro - physical activity monitor processing [Internet]. GitHub. 2021. https://github.com/MRC-Epid/pampro

15. Van Hees VT, Fang Z, Langford J, Assah F, Mohammad A, Da Silva IC, et al. Autocalibration of accelerometer data for free-living physical activity assessment using local gravity and temperature: an evaluation on four continents. J Appl Physiol. 2014;117(7):738–44.

16. van Hees VT, Renström F, Wright A, Gradmark A, Catt M, Chen KY, et al. Estimation of daily energy expenditure in pregnant and non-pregnant women using a wrist-worn tri-axial accelerometer. PLoS One. 2011;6(7):e22922.

17. Koivula RW, Atabaki-Pasdar N, Giordano GN, White T, Adamski J, Bell JD, et al. The role of physical activity in metabolic homeostasis before and after the onset of type 2 diabetes: an IMI DIRECT study. Diabetologia. 2020;63(4):744–56.

18. Hildebrand M, Van Hees VT, Hansen BH, Ekelund U. Age group comparability of raw accelerometer output from wrist-and hip-worn monitors. Med Sci Sports Exerc. 2014;46(9):1816–24.

19. Edwardson CL, Rowlands AV, Bunnewell S, Sanders JP, Esliger D, Gorely T, et al. Accuracy of posture allocation algorithms for thigh-and waist-worn accelerometers. Med Sci Sports Exerc. 2016;48(6):1085–90.

20. Hartley P, Keevil VL, Westgate K, White T, Brage S, Romero-Ortuno R, et al. Using accelerometers to measure physical activity in older patients admitted to hospital. Current Gerontology and Geriatrics Research. 2018;2018.

21. Brage S, Westgate K, Wijndaele K, Godinho J, Griffin S, Wareham N, editors. Evaluation of a method for minimising diurnal information bias in objective sensor data. ICAMPAM; 2013; Amherst, MA.

22. World Health Organization. Obesity: preventing and managing the global epidemic. World Health Organization Technical Report Series. 2000;894:1–253.

23. Fahrenberg J, Foerster F, Smeja M, Müller W. Assessment of posture and motion by multichannel piezoresistive accelerometer recordings. Psychophysiology. 1997;34(5):607–12.

24. Zhang K, Werner P, Sun M, Pi[Sunyer FX, Boozer CN. Measurement of human daily physical activity. Obes Res. 2003;11(1):33–40.

25. Bussmann JBJ, Tulen JHM, van Herel ECG, Stam HJ. Quantification of physical activities by means of ambulatory accelerometry: a validation study. Psychophysiology. 1998;35(5):488–96.

26. De Vries SI, Garre FG, Engbers LH, Hildebrandt VH, Van Buuren S. Evaluation of neural networks to identify types of activity using accelerometers. Med Sci Sports Exerc. 2011;43(1):101–7.

27. Skotte J, Korshøj M, Kristiansen J, Hanisch C, Holtermann A. Detection of physical activity types using triaxial accelerometers. Journal of Physical Activity and Health. 2014;11(1):76–84.

28. Grant PM, Ryan CG, Tigbe WW, Granat MH. The validation of a novel activity monitor in the measurement of posture and motion during everyday activities. Br J Sports Med. 2006;40(12):992–7.

29. Kozey-Keadle S, Libertine A, Lyden K, Staudenmayer J, Freedson PS. Validation of wearable monitors for assessing sedentary behavior. Med Sci Sports Exerc. 2011;43(8):1561–7.

30. Edwardson CL, Rowlands AV, Bunnewell S, Sanders JP, Esliger D, Gorely T, et al. Accuracy of posture allocation algorithms for thigh-and waist-worn accelerometers. Med Sci Sports Exerc. 2016;48(6):1085–90.

31. Stemland I, Ingebrigtsen J, Christiansen CS, Jensen BR, Hanisch C, Skotte J, et al. Validity of the Acti4 method for detection of physical activity types in free-living settings: comparison with video analysis. Ergonomics. 2015;58(6):953–65.

32. Lunde LK, Koch M, Knardahl S, Veiersted KB. Associations of objectively measured sitting and standing with low-back pain intensity: a 6-month follow-up of construction and healthcare workers. Scand J Work Environ Health. 2017;43(3):269–78.

33. Hallman DM, Birk Jørgensen M, Holtermann A. On the health paradox of occupational and leisure-time physical activity using objective measurements: effects on autonomic imbalance. PLoS One. 2017;12(5):e0177042.

34. Reinsve Ø. Data analytics for hunt: Recognition of physical activity on sensor data streams: NTNU; 2018.

35. Chastin SFM, Van Cauwenberg J, Maenhout L, Cardon G, Lambert EV, Van Dyck D. Inequality in physical activity, global trends by income inequality and gender in adults. International Journal of Behavioral Nutrition and Physical Activity. 2020;17(1):1–8.

36. Guthold R, Stevens GA, Riley LM, Bull FC. Worldwide trends in insufficient physical activity from 2001 to 2016: a pooled analysis of 358 population-based surveys with 1.9 million participants. The Lancet Global Health. 2018;6(10):e1077–e86.

37. Howitt C, Brage S, Hambleton IR, Westgate K, Samuels TA, Rose AMC, et al. A cross-sectional study of physical activity and sedentary behaviours in a Caribbean population: combining objective and questionnaire data to guide future interventions. BMC Public Health. 2016;16(1):1–12.

38. Laverty AA, Palladino R, Lee JT, Millett C. Associations between active travel and weight, blood pressure and diabetes in six middle income countries: a cross-sectional study in older adults. International Journal of Behavioral Nutrition and Physical Activity. 2015;12(1):1–11.

39. Maimela E, Alberts M, Modjadji SEP, Choma SSR, Dikotope SA, Ntuli TS, et al. The prevalence and determinants of chronic non-communicable disease risk factors amongst adults in the Dikgale health demographic and surveillance system (HDSS) site, Limpopo Province of South Africa. PLoS One. 2016;11(2):e0147926.

40. Gradidge PJL, Crowther NJ, Chirwa ED, Norris SA, Micklesfield LK. Patterns, levels and correlates of self-reported physical activity in urban black Soweto women. BMC Public Health. 2014;14(1):934.

41. Prioreschi A, Wrottesley SV, Norris SA. Physical Activity Levels, Food Insecurity and Dietary Behaviours in Women from Soweto, South Africa. J Community Health. 2021;46(1):156–64.

42. O’donoghue G, Perchoux C, Mensah K, Lakerveld J, Van Der Ploeg H, Bernaards C, et al. A systematic review of correlates of sedentary behaviour in adults aged 18–65 years: a socio-ecological approach. BMC Public Health. 2016;16(1):1–25.

43. Patterson R, McNamara E, Tainio M, de Sá TH, Smith AD, Sharp SJ, et al. Sedentary behaviour and risk of all-cause, cardiovascular and cancer mortality, and incident type 2 diabetes: a systematic review and dose response meta-analysis. Eur J Epidemiol. 2018;33(9):811–29.

44. van der Ploeg HP, Chey T, Ding D, Chau JY, Stamatakis E, Bauman AE. Standing time and all-cause mortality in a large cohort of Australian adults. Prev Med. 2014;69:187–91.

45. Chastin SFM, Dontje ML, Skelton DA, Čukić I, Shaw RJ, Gill JMR, et al. Systematic comparative validation of self-report measures of sedentary time against an objective measure of postural sitting (activPAL). International Journal of Behavioral Nutrition and Physical Activity. 2018;15(1):1–12.

46. Ekelund U, Kolle E, Steene-Johannessen J, Dalene KE, Nilsen AKO, Anderssen SA, et al. Objectively measured sedentary time and physical activity and associations with body weight gain: does body weight determine a decline in moderate and vigorous intensity physical activity? Int J Obes. 2017;41(12):1769–74.

47. Ekelund U, Brage S, Besson S, Sharp S, Wareham NJ. Time spent being sedentary and weight gain in healthy adults: reverse or bidirectional causality? The American Journal of Clinical Nutrition. 2008;88(3):612–7.

48. Golubic R, Ekelund U, Wijndaele K, Luben R, Khaw KT, Wareham NJ, et al. Rate of weight gain predicts change in physical activity levels: a longitudinal analysis of the EPIC-Norfolk cohort. Int J Obes. 2013;37(3):404–9.

49. Micklesfield LK, Munthali RJ, Prioreschi A, Said-Mohamed R, Van Heerden A, Tollman S, et al. Understanding the relationship between socio-economic status, physical activity and sedentary behaviour, and adiposity in young adult South African women using structural equation modelling. Int J Environ Res Public Health. 2017;14(10):1271.

50. Micklesfield LK, Pedro TM, Kahn K, Kinsman J, Pettifor JM, Tollman S, et al. Physical activity and sedentary behavior among adolescents in rural South Africa: levels, patterns and correlates. BMC Public Health. 2014;14(1):1–10.

51. Mlangeni L, Makola L, Naidoo I, Chibi B, Sokhela Z, Silimfe Z, et al. Factors associated with physical activity in South Africa: evidence from a National Population Based Survey. The Open Public Health Journal. 2018;11(1).

52. Tomaz SA, Davies JI, Micklesfield LK, Wade AN, Kahn K, Tollman SM, et al. Self-Reported Physical Activity in Middle-Aged and Older Adults in Rural South Africa: Levels and Correlates. Int J Environ Res Public Health. 2020;17(17):6325–37.

53. Rae DE, Pienaar PR, Henst RHP, Roden LC, Goedecke JH. Associations between long self-reported sleep, obesity and insulin resistance in a cohort of premenopausal black and white South African women. Sleep Health. 2018;4(6):558–64.

