## Supplementary figures and images for "Physical activity and posture profile of a South African cohort of middle-aged men and women as determined by integrated hip and thigh accelerometry"

### Supplementary Figure S1

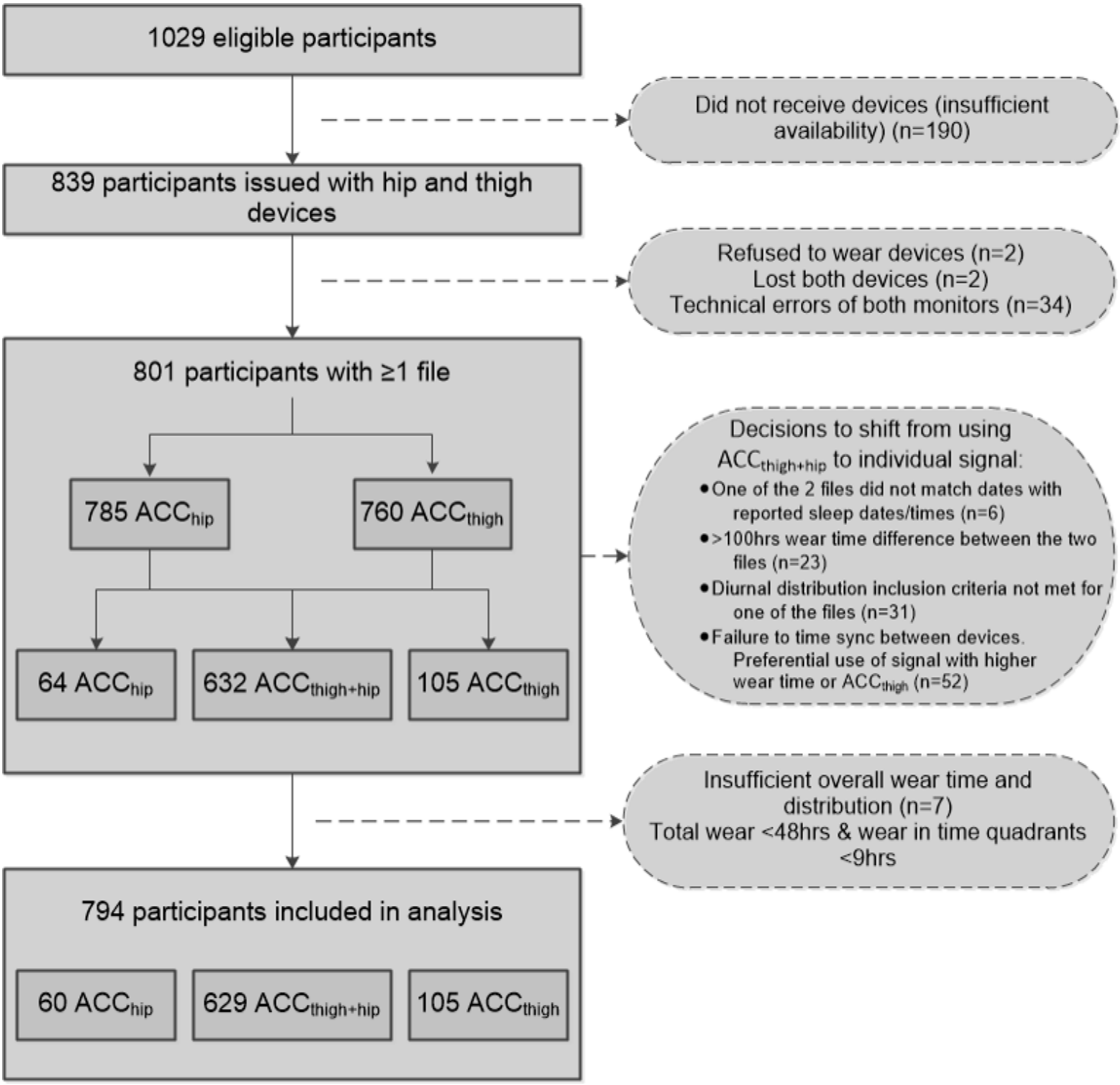
