## Supplementary Table S1 for "Physical activity and posture profile of a South African cohort of middle-aged men and women as determined by integrated hip and thigh accelerometry"

**Additional file 2 - Table S1:** Descriptive characteristics of sub-sample included and excluded from the current study.

|  | **Men (n=502)** | | | | | **Women (n=527)** | | | | |
| --- | --- | --- | --- | --- | --- | --- | --- | --- | --- | --- |
|  | **Included (n=437)** | | **Excluded (n=65)** | | **p-value** | **Included (n=357)** | | **Excluded (n=170)** | | **p-value** |
| **Age (years)** | 53.6 | 6.2 | 53.5 | 5.8 | 0.961 | 53.9 | 5.8 | 55.4 | 6.1 | 0.008 |
| **BMI (kg/m^2^)** | 25.6 | 6.0 | 25.8 | 6.2 | 0.757 | 33.5 | 6.8 | 33.5 | 7.9 | 0.961 |
| **Waist circumference (cm)** | 94.2 | 15.3 | 95.8 | 15.8 | 0.430 | 96.4 | 13.6 | 97.3 | 13.9 | 0.487 |
| **Currently employed** | 178 | 40.7 | 23 | 36.5 | 0.523 | 139 | 38.9 | 60 | 36.8 | 0.644 |
| **Married** | 241 | 55.1 | 28 | 44.4 | 0.111 | 148 | 41.4 | 68 | 41.7 | 0.955 |
| **Current smoker** | 220 | 50.3 | 27 | 42.8 | 0.267 | 22 | 6.2 | 10 | 6.2 | 0.996 |
| **Education Level** |  |  |  |  | 0.130 |  |  |  |  | 0.893 |
| No formal/elementary | 4 | 0.9 | 0 | 0 |  | 3 | 0.8 | 2 | 1.2 |  |
| Secondary | 350 | 80.1 | 57 | 90.5 |  | 312 | 87.4 | 143 | 87.7 |  |
| Tertiary | 83 | 19 | 6 | 9.5 |  | 42 | 11.8 | 18 | 11 |  |
| **Alcohol intake** |  |  |  |  | 0.415 |  |  |  |  | 0.512 |
| Never | 116 | 26.5 | 17 | 27 |  | 248 | 69.5 | 118 | 72.8 |  |
| Sometimes | 176 | 40.3 | 30 | 47.6 |  | 93 | 26 | 35 | 21.6 |  |
| Often | 145 | 33.2 | 16 | 25.4 |  | 16 | 4.5 | 9 | 5.6 |  |

Participant numbers in excluded category may differ for different variables depending on availability of data.
