## Supplementary Table S2 for "Physical activity and posture profile of a South African cohort of middle-aged men and women as determined by integrated hip and thigh accelerometry"

**Additional file 3 - Table S2:** Multivariable analysis of physical activity volume and time in MVPA using hip-worn accelerometer signal (ACC_hip_) only

|  | **ENMO ACC_hip_** | | | | **MVPA ACC_hip_** | | | |
| --- | --- | --- | --- | --- | --- | --- | --- | --- |
|  | **MEN** | | **WOMEN** | | **MEN** | | **WOMEN** | |
| **Age (years)** |  |  |  |  |  |  |  |  |
| 41 - 49 | Reference | | Reference | | Reference | | Reference | |
| 50 - 59 | -1.1** | -2.1 - -0.1 | -0.6 | -1.3 - 0.2 | -6.9 | -15.4 - 1.6 | -3.7 | -9.4 - 2 |
| 60 - 72 | -3.1*** | -4.3 - -1.8 | -1.7*** | -2.7 - -0.6 | -16.9*** | -27.7 - -6.1 | -13.6*** | -21.5 - -5.7 |
| **BMI (kg/m2)** |  |  |  |  |  |  |  |  |
| <18.5 | -1.9** | -3.4 - -0.3 | -3.9* | -8.2 - 0.4 | -8 | -21.6 - 5.5 | -23.7 | -55.4 - 7.9 |
| 18.5-24.9 | Reference | | Reference | | Reference | | Reference | |
| 25 - 29.9 | -1.7*** | -2.7 - -0.6 | -0.4 | -1.6 - 0.9 | -18.6*** | -27.8 - -9.5 | 1.3 | -7.8 - 10.4 |
| 30 - 34.9 | -2.5*** | -3.8 - -1.2 | -0.6 | -1.8 - 0.6 | -23.3*** | -34.7 - -12 | -0.2 | -8.9 - 8.5 |
| 35 - 39.9 | -1.9* | -4.1 - 0.3 | -1.4** | -2.6 - -0.1 | -9.8 | -28.8 - 9.2 | -7 | -16.1 - 2 |
| >=40 | -3.4** | -6.3 - -0.5 | -1.6** | -2.9 - -0.3 | -43.1*** | -68.1 - -18.2 | -7.7 | -17.3 - 1.9 |
| **HIV Status** |  |  |  |  |  |  |  |  |
| HIV uninfected | Reference | | Reference | | Reference | | Reference | |
| HIV infected | -0.5 | -1.6 - 0.6 | -0.7* | -1.6 - 0.1 | -2.1 | -11.7 - 7.4 | -1.9 | -8.3 - 4.5 |
| **Education Level** |  |  |  |  |  |  |  |  |
| No formal/elementary | Reference | | Reference | | Reference | | Reference | |
| Secondary | 1.5** | 0.1 - 2.8 | -0.2 | -1.5 - 1.1 | 12.4** | 0.5 - 24.3 | -0.1 | -9.6 - 9.5 |
| Tertiary | 1.1 | -0.6 - 2.8 | -1.3 | -2.9 - 0.3 | 9.4 | -5.1 - 23.9 | -8.9 | -20.8 - 3 |
| **SES (Asset count)** |  |  |  |  |  |  |  |  |
| Asset count in household^†^ | -0.4*** | -0.5 - -0.2 | -0.2** | -0.3 - 0 | -4.9*** | -6.6 - -3.2 | -2.9*** | -4.1 - -1.8 |
| **Seasonality** |  |  |  |  |  |  |  |  |
| SPRING | 0.1 | -0.6 - 0.8 | -0.2 | -0.7 - 0.2 | 0.2 | -5.7 - 6.1 | -1.2 | -4.3 - 2 |
| WINTER | 0 | -0.7 - 0.7 | -0.5* | -1 - 0 | 0.8 | -5 - 6.6 | -1.7 | -5.4 - 2 |
| Constant^†^ | 13.7*** | 12.0 - 15.3 | 12.2*** | 10.4 - 14.0 | 64.0*** | 49.9 - 78.0 | 40.0*** | 26.5 - 53.5 |

*** p<0.01, ** p<0.05, * p<0.1

^†^Centred at 9 assets
